# SARS-CoV-2 vaccine effectiveness and breakthrough infections in maintenance dialysis patients

**DOI:** 10.1101/2021.12.20.21268124

**Authors:** Harold J. Manley, Gideon N. Aweh, Caroline M. Hsu, Daniel E. Weiner, Dana Miskulin, Antonia M. Harford, Doug Johnson, Eduardo K. Lacson

**Affiliations:** Dialysis Clinic Inc., Nashville, TN; Division of Nephrology, Tufts Medical Center, Boston, Massachusetts

**Keywords:** Dialysis, COVID-19 infection, breakthrough, antibody levels

## Abstract

**Background:** SARS-CoV-2 vaccine effectiveness during the Delta period and immunogenicity threshold associated with protection against COVID-19 related hospitalization or death in the dialysis population is unknown.

**Methods:** A retrospective, observational study assessed SARS-CoV-2 vaccine effectiveness and immunogenicity threshold in all adult maintenance dialysis patients without COVID-19 history treated between February 1 and October 2, 2021. All COVID-19 infections, composite of hospitalization or death following COVID-19 and available SARS-CoV-2 anti-spike immunoglobulin (Ig) G values were extracted from electronic medical record. COVID-19 cases per 10,000 days at risk and vaccine effectiveness during pre-Delta and Delta periods were determined.

**Results:** Of 15,718 patients receiving dialysis during the study period, 11,191 (71%) were fully vaccinated, 733 (5%) were partially vaccinated and 3,794 (24%) were unvaccinated. 967 COVID-19 were cases identified: 511 (53%) occurred in unvaccinated patients and 579 (60%) occurred during the Delta period. COVID-19 related hospitalization or death was less likely among vaccinated versus unvaccinated patients for all vaccines (adjusted HR 0.19 [0.12, 0.30]) and for BNT162b2/Pfizer, mRNA-1273/Moderna, and Ad26.COV2.S/Janssen (adjusted HR=0.25 [0.16, 0.40], 0.14 [0.08, 0.22], and 0.34 [0.17, 0.68] respectively). Among those with anti-spike IgG levels, those with IgG level ≥ 7 had significantly lower risk of a COVID-19 diagnosis (HR=0.25 [0.15, 0.42]) and none experienced a COVID-related hospitalization or death.

**Conclusions:** Among maintenance dialysis patients, SARS-CoV-2 vaccination was associated with a lower risk of COVID-19 diagnosis and associated hospitalization or death. Among vaccinated patients, low anti-spike IgG level is associated with worse COVID-19 related outcomes.

**Significance Statement:** SARS-CoV-2 vaccine effectiveness and association between antibody levels and clinical outcomes in maintenance dialysis patients is not known. Between February 1 and October 2, 2021, vaccine effectiveness was 85% against COVID-19 infection and 81% against composite of COVID-related hospitalization or death. COVID-19 case rates and severe outcomes were higher during the Delta dominant period (June 27-October 2, 2021). Increasing time (weeks) since full vaccination status was associated with increased risk for COVID-19 related hospitalization or death. Anti-spike IgG level ≥ 7 had lower risk of a COVID-19 diagnosis and no COVID-related hospitalization or death. Our findings supports utilization of SARS-CoV-2 vaccination and suggests that monitoring SARS-CoV-2 antibody levels and administering additional vaccine doses to maintain adequate immunity will be beneficial.

## Introduction

Maintenance dialysis patients experience significant COVID-19–associated morbidity and mortality.^1, 2^ Vaccines are an effective tool for combatting COVID-19, both in the general population and, likely, among patients dependent on maintenance dialysis, and studies indicate that two doses of a SARS-CoV-2 mRNA vaccine elicit a seroresponse in most (>90%) maintenance dialysis patients, albeit with lower levels than in the general population.^3-5^

Since June 26, 2021, the Delta variant has become the dominant SARS-CoV-2 strain in the United States.^6^ In addition to increased virulence, the increased breakthrough rate in fully vaccinated patients during the Delta variant dominant period is also likely due to waning antibody concentrations. In the general population, Tartof et al reported a decrease of protection offered by the BNT162b2/Pfizer vaccine from 93% at baseline to 53% after at least 4 months.^7^ We previously reported that 10-15% of dialysis patients did not respond to two doses of an mRNA vaccine; additionally, among initial responders to vaccine, more than half experience waning immunity by 4-6 months, particularly those whose initial response was lesser.^5, 8^

The impact of lesser initial vaccine response and subsequent waning antibody levels on clinical outcomes among maintenance dialysis patients is not known. Additionally, the impact of the Delta variant on vaccine effectiveness in this population is unknown. Accordingly, we describe the incidence of COVID-19 diagnoses and COVID-19 related hospitalization or death among unvaccinated, partially vaccinated and fully vaccinated adult dialysis patients treated by a national dialysis provider within the US between February 1 and October 2, 2021, evaluating both the pre-Delta and Delta dominant periods. Additionally, among the subset of vaccinated patients with available SARS-CoV-2 vaccine immunoglobulin G spike antibody (anti-spike IgG) titers who were diagnosed with ‘breakthrough’ COVID-19, we explore the association between antibody levels and clinical outcomes.

## Methods

### Study Population

Dialysis Clinic, Incorporated (DCI) is the largest non-profit provider of kidney care and dialysis care in the United States, operating 260 outpatient dialysis clinics in 29 states, 20 chronic kidney disease clinics in 5 states, and 3 organ procurement organizations in 3 states. All adult (age ≥18 years) maintenance dialysis patients treated between February 1 and October 02, 2021 in DCI clinics contributed time-at-risk, excluding patients with known COVID-19 diagnosis prior to February 1, 2021, transient patients (e.g. visiting from a non-DCI clinic) and/or patients treated for acute kidney injury who were not certified as end-stage renal disease (ESRD).

### Screening for COVID-19

All maintenance dialysis patients treated at DCI outpatient facilities, including home dialysis and in-center dialysis patients, are screened at each clinic encounter for COVID-19 symptoms and any known exposure to infected person(s). These symptoms include fever (≥100 degrees), sore throat, new or worsening cough or shortness of breath, and loss of taste or smell. The vast majority of maintenance dialysis patients (>85%) are treated by in-center dialysis and are screened before each treatment, most often thrice weekly, while home dialysis patients typically are screened once or twice each month during visits to the dialysis facility. Any patient who screens positive is classified as a “Patient Under Investigation” and is tested for SARS-CoV-2 either locally or through the DCI laboratory. Most dialysis patients have multiple comorbid illnesses and have frequent contact with the healthcare system where they are tested even when asymptomatic. Similarly, many patients who are residents of congregate home settings, such as nursing homes, are frequently screened and tested as well. The diagnosis of COVID-19 is entered into DCI’s electronic health record (EHR), often based on a report of a positive SARS-CoV-2 test result, which if performed via the DCI laboratory, utilizes a reverse transcriptase–polymerase chain reaction (RT-PCR) Cobas SARS-CoV-2 Assay [Roche Diagnostics].

### SARS-CoV-2 Antibody Testing

The DCI central laboratory assesses serum anti-spike IgG antibody against the receptor binding domain of the S1 subunit of SARS-CoV-2 spike antigen using a US-FDA-EUA-approved chemiluminescent assay (ADVIA Centaur® XP/XPT COV2G). This semi-quantitative assay has a range between 0 and ≥20. The manufacturer defined threshold for a minimum antibody detection level is ≥1.^9^ This test is available for physicians to order either one time or as part of a clinical testing protocol, but is not measured in all maintenance dialysis patients routinely, per present CDC guidelines.^10^ The clinical testing protocol follows monthly anti-spike IgG titers using residual blood from routine monthly lab draws until the level is <1 for two consecutive months or for up to 12 consecutive months.

### Outcomes

New COVID-19 diagnoses were documented based on screening as described above, or from contacts with other healthcare settings, such as hospitals and emergency department visits or clinic visits. All new COVID-19 diagnoses that occurred during the study period were each assigned to the appropriate vaccination status. For all breakthrough cases, defined as a COVID-19 diagnosis in a patient deemed fully vaccinated, the clinic was contacted to verify the reason why the patient was tested for SARS-CoV-2: for exposure to a known COVID-19 infected person <u>or</u> for symptoms/signs consistent with COVID-19 <u>or</u> as required by a non-dialysis clinic/hospital protocol prior to providing a COVID-unrelated service/procedure (e.g. vascular access surgery) <u>or</u> as part of protocolized screening (e.g. nursing home). COVID-19 related hospitalizations or deaths were defined by documented primary diagnosis for the episode of care as COVID-19 (ICD-10 code U01.7).

COVID-19 cases, hospitalizations and deaths were identified throughout the study period, and further subdivided into pre-COVID-19 delta variant (February 1, 2021 - June 25, 2021) and COVID-19 delta variant dominant periods (June 26, 2021 - October 02, 2021). The end of the follow up period to identify any patient diagnosed with COVID-19 on October 2, 2021 or earlier as having a COVID-19 related hospitalization or death within 30 days of diagnosis was November 02, 2021.

### Data Extraction and Analysis

All patient demographic and clinical variables used in this analyses were retrospectively obtained from the DCI EHR. These variables include: SARS-CoV-2 vaccine name and dates administered, age, sex, race (Black, White, Native American, Asian/Pacific Islander, Other/Unknown), ethnicity (Hispanic or Non-Hispanic), US state and county of residence, congregate living status (e.g., nursing home, long term care facility), modality (in-center hemodialysis, home hemodialysis, peritoneal dialysis), date of ESRD, body mass index, dialysis dose delivered (Kt/V), serum albumin, hepatitis B surface antibody, immunosuppression (immune-modulating medications, prior transplant, immunodeficiency disorder), substance abuse disorder (tobacco, alcohol or drug), and other comorbid conditions. For this study, any SARS-CoV-2 infection occurring more than 14 days after completing SARS-CoV-2 vaccination was deemed a breakthrough COVID-19 case even if asymptomatic. Data integrity checks for COVID-19 documentation are performed weekly. These include comparing consistency of all available concurrent documentation, including: *de novo* ICD-10 COVID-19 diagnoses within the problem list, Patient Under Investigation status during symptom screening every visit, new COVID-19 diagnosis screening indicator during every visit, as well as any lab report indicating a positive test for SARS-CoV-2. Inconsistencies detected are referred to corporate nurses who directly communicate with individual clinic staff for resolution.

### Exposure and Time at Risk

Each eligible patient who received treatment for at least one day during the study period contributed days-at-risk. Patients could move from being unvaccinated (defined by the CDC to include the period extending up to 13 days after the first vaccine dose)^3^ to partially vaccinated (for mRNA vaccines only; defined by the CDC as the period starting 14 days after the first mRNA vaccine dose up to 13 days after the 2^nd^ mRNA vaccine dose) to fully vaccinated (≥14 days after completing the manufacturer recommended final dose). Patients could contribute days-at-risk for each applicable category as long as they remained SARS-CoV-2 uninfected.

Case rates per 10,000 days at-risk and the composite outcome of COVID-related hospitalization or death for each vaccine status, and, in additional analyses among those fully vaccinated, for each vaccine type, were compared using a time dependent Cox proportional hazard models with unvaccinated patients as the reference group. The statistical model was subsequently adjusted for all baseline patient characteristics that differed at baseline among vaccination status within each time period (**Table 1**). Each patient’s state and county of residence was also used to adjust for geotemporal variability in the intensity of the epidemic and likelihood to follow mask wearing, social distancing and vaccine recommendations by incorporating daily county-level COVID-19 case rates and the percentage of Biden voters during 2020 US Presidential election, respectively.^11-13^ Vaccine effectiveness and 95% CIs were calculated using the following: (1 – HR) x 100.^14^ We also determined case rates and vaccine effectiveness in a subset of patients vaccinated during the Delta variant dominant period.

**Table 1.** Patient characteristics over entire study period (February 1-October 2, 2021)

| **Demographics** | **All Patients**<br>**(N=15,718)** | **Fully**<br>**Vaccinated**<br>**(N=11,191)** | **Partially**<br>**Vaccinated**<br>**(N=733)** | **Unvaccinated**<br>**(N=3,794)** | **P-value** |
| --- | --- | --- | --- | --- | --- |
| **Age years** (mean ± SD) | 63 ± 15 | 65 ± 14 | 61 ± 15 | 58 ± 16 | <0.0001 |
| **Age ≥ 65 years** | 7,572 (48) | 5,877 (52) | 309 (42) | 1,386 (37) | <0.0001 |
| **Age – Decade** |  |  |  |  | <0.0001 |
| <55 | 4,300 (27) | 2,540 (23) | 243 (33) | 1,517 (40) |  |
| 55-64 | 3,846 (24) | 2,774 (25) | 181 (25) | 891 (23) |  |
| 65-74 | 4,294 (27) | 3,299 (29) | 190 (26) | 805 (21) |  |
| 75+ | 3,278 (21) | 2,578 (23) | 119 (16) | 581 (15) |  |
| **Female** | 6,653 (42) | 4,578 (41) | 329 (45) | 1,746 (46) | <0.0001 |
| **Race** |  |  |  |  | <0.0001 |
| White | 7,370 (47) | 5,325 (48) | 331 (45) | 1,714 (45) |  |
| Black | 5,605 (36) | 3872 (35) | 280 (38) | 1,453 (38) |  |
| Native American | 357 (2) | 286 (3) | 14 (2) | 57 (2) |  |
| Asian/Pacific Islander | 491 (3) | 398 (4) | 18 (2) | 75 (2) |  |
| Other/Unknown | 1,895 (12) | 1,310 (12) | 90 (12) | 495 (13) |  |
| **Hispanic** | 918 (6) | 675 (6) | 37 (5) | 206 (5) | 0.25 |
| **Vintage months**<br>(mean ± SD) | 45 ± 56 | 46 ± 56 | 40 ± 55 | 43 ± 56 | 0.001 |
| **Body Mass Index (kg/m<sup>2</sup>)**<br>(mean ± SD) | 29 ± 8 | 29 ± 7 | 29 ± 8 | 28 ± 8 | 0.15 |
| Congregate Living <sup>a</sup> | 2,005 (13) | 1,398 (12) | 134(18) | 473 (12) | <0.0001 |
| **Home Dialysis** | 2,075 (13) | 1,526 (14) | 75 (10) | 474 (12) | 0.0107 |
| Peritoneal Dialysis | 1,906 (12) | 1397 (12) | 73 (10) | 436 (12) |  |
| Home Hemodialysis | 169 (1) | 129 (1) | 2 (0) | 38 (1) |  |
| Adequate Dialysis Dose <sup>b</sup> | 11,318 (72) | 8,393 (75) | 533 (73) | 2,392 (63) | <0.0001 |
| Serum Albumin (g/dl)<br>(mean ± SD) | 3.8 ± 0.5 | 3.9 ± 0.5 | 3.8 ± 0.5 | 3.8 ± 0.5 | <0.0001 |
| History Hepatitis B Positive<br><sup>c</sup> | 6,797 (43) | 6,440 (58) | 261 (36) | 96 (3) | 0.007 |
| **Potential**<br>**Immunosuppression** | 3,098 (20) | 2,205 (20) | 147 (20) | 746 (20) | 0.97 |
| Immune-modulating Meds | 2,389 (15) | 1,703 (15) | 117 (16) | 569 (15) | 0.80 |
| Prior Transplant | 996 (6) | 744(7) | 40 (5) | 212 (6) | 0.03 |
| Immunodeficiency<br>Disorder | 1,783 (11) | 1,297 (12) | 81 (11) | 405 (11) | 0.30 |
| Disability | 725 (5) | 508 (5) | 48 (7) | 169 (4) | 0.04 |
| Tobacco Use | 2,528 (16) | 1,783 (16) | 132 (18) | 613 (16) | 0.33 |
| Alcohol Abuse Disorder | 1,583 (10) | 1,151 (10) | 88 (12) | 344 (9) | 0.02 |
| Drug Abuse Disorder | 797 (5) | 530 (5) | 41 (6) | 226 (6) | 0.01 |
| Number of Comorbidities<br>(mean ± SD) | 3.0 ± 1.8 | 3.0 ± 1.8 | 3.2 ± 1.8 | 2.7 ± 1.8 | <0.0001 |
| Diabetes Mellitus | 9,181 (59) | 6,704 (60) | 468 (64) | 2,009 (53) | <0.0001 |
| Hypertension | 12,867 (82) | 9,293 (83) | 609 (83) | 2,965 (78) | <0.0001 |
| Congestive Heart Failure | 3,480 (22) | 2,440 (22) | 190 (26) | 850 (22) | 0.03 |
| COPD | 2,418 (15) | 1,751 (16) | 117 (16) | 550 (15) | 0.22 |
| Stroke/Cerebrovascular<br>Disorder | 1,464 (9) | 1,064 (10) | 79 (11) | 321 (8) | 0.06 |
| Peripheral Vascular<br>Disease | 2,059 (13) | 1,515 (14) | 113 (15) | 431 (11) | 0.001 |
| Thyroid Disorder | 2,393 (15) | 1,800 (16) | 95 (13) | 498 (13) | <0.0001 |
| History of Cancer | 1,521 (10) | 1,152 (10) | 63 (9) | 306 (8) | 0.0002 |
| Percent Biden Voters<br>(median [IQR]) | 49 (25) | 49 (26) | 48 (28) | 47 (26) | <0.0001 |

<sup>a</sup> Residing in nursing home or long-term care facility
<sup>b</sup> Adequate dialysis defined by hemodialysis single pool Kt/V ≥ 1.2 or peritoneal dialysis weekly Kt/V ≥ 1.7
<sup>c</sup> Hepatitis B seroimmunity defined as hepatitis B surface antibody ≥ 10 mIU/mL
Results presented as count (N) and percent (%) unless otherwise specified. Patients are grouped based on their vaccination status at the end of follow-up. Unvaccinated includes patients who never received a vaccine or recipients of a single dose of a vaccine within 14 days of vaccine receipt; partially vaccinated includes patients who were ≥ 14 days after the first mRNA vaccine dose but <14 days after the 2nd mRNA vaccine dose; fully vaccinated patients includes all patients ≥ 14 days after the last vaccine dose. For the table, categories are mutually exclusive.
IQR = interquartile range; SD = standard deviation; COPD = Chronic Obstructive Pulmonary Disease

Among patients with breakthrough COVID-19 diagnoses, we evaluated the subset with available anti-spike IgG titer results prior to the COVID-19 diagnosis. Results subsequently were de-identified and aggregated, and the association between the most proximate anti-spike IgG titer result and clinical outcomes was evaluated descriptively. Case rates per 10,000 days and adjusted hazard ratios for COVID-19 infection and composite for COVID-related hospitalization or death were calculated for anti-spike IgG values <1 (versus ≥1), < 2 (versus ≥2), <7 (versus ≥7) and <10 (versus ≥10). We selected the various cut-points to compare outcomes in those with undetectable levels of < 1 (reported by assay manufacturer) or < 2 (internal DCI laboratory validation), at assay threshold one index value above where COVID-related hospitalization or death not reported (anti-spike IgG value ≥7) and assay level recently reported as having higher odds for breakthrough infection (anti-spike IgG < 10).^15^

This study was reviewed and approved by the WCG IRB Work Order 1-1456342-1. Statistical analyses were performed using SAS v9.4.

## Results

Among 15,718 maintenance dialysis patients at DCI facilities during the study period, 11,191 (71%) were fully vaccinated by October 2, 2021, 6,156 (55%) with mRNA-1273/Moderna, 4,490 (40%) with BNT162b2/Pfizer, and 545 (5%) with Ad26.COV2.S/Janssen). An additional 733 (5%) patients were partially vaccinated, 426 (58%) with mRNA-1273/Moderna and 307 (42%) with BNT162b2/Pfizer, while 3,794 (24%) were unvaccinated. Patients mean age (years) and vintage (months) were 63±15 years old and 45 ± 56 months, respectively. Majority (87%) patients were receiving in-center hemodialysis, 59% had diabetes and 20% were considered immunocompromised (**Table 1**).

### Vaccination status and Vaccine Effectiveness against COVID-19 infection

There were 967 documented COVID-19 cases, with 511 (53%) occurring in unvaccinated patients and 579 (60%) occurring during the Delta dominant period. COVID-19 rates per 10,000 patient days and vaccine effectiveness for each time period between February 1 and October 2, 2021 are shown in **Table 2**. There were 362 (37%) cases that occurred among those considered fully vaccinated, with most of these breakthrough cases (n=335; 93%) occurring during the Delta dominant period; 92 (25%) of patients with breakthrough cases met CDC criteria for immunosuppressed. The median (IQR) time from being considered fully vaccinated to COVID-19 diagnosis was 133 (105 -156) days (**Fig S1 Panel A**). There was no difference in median days to breakthrough infection among vaccines: mRNA-1273/Moderna 137 (110, 158) days; BNT162b2/Pfizer 131 (102, 158) days; Ad26.COV2.S/Janssen 129 (100, 145) days; p=0.58 (**Figure S1 Panel B**).

**Table 2.** COVID-19 infection rates per 10,000 patient days and vaccine effectiveness between February 1 and October 2, 2021.

|  | **Last**<br>**Vaccination**<br>**Status\*** | **Person**<br>**Days at**<br>**Risk** | **COVID-19**<br>**diagnoses** | **Rate per**<br>**10,000**<br>**days** | **Adjusted Hazard**<br>**Ratio (95% CI)** | **Vaccine**<br>**Effectiveness**<br>**(95% CI)** |
| --- | --- | --- | --- | --- | --- | --- |
| February 1 – October 2, 2021 |  |  |  |  |  |  |
| Unvaccinated | 3,794 | 1,349,022 | 511 | 3.8 | Reference | Reference |
| Partially Vaccinated | 733 | 345,742 | 94 | 2.7 | 0.87 (0.69, 1.09) | 13% |
| Fully Vaccinated | 11,191 | 1,810,193 | 362 | 2.0 | 0.15 (0.11, 0.20) | 85% |
| BNT162b2/Pfizer | 4,490 | 718,411 | 171 | 2.4 | 0.17 (0.12, 0.24) | 83% |
| mRNA-1273/Moderna | 6,156 | 1,009,602 | 160 | 1.6 | 0.12 (0.08, 0.16) | 88% |
| Ad26.COV2.S/Janssen | 545 | 82,180 | 31 | 3.8 | 0.27 (0.17, 0.42) | 73% |
| February 1- June 26, 2021 – pre-Delta variant period |  |  |  |  |  |  |
| Unvaccinated | 3,809 | 926,858 | 291 | 3.1 | Reference | Reference |
| Partially Vaccinated | 664 | 278,500 | 65 | 2.3 | 0.88 (0.66, 1.17) | 12% |
| Fully Vaccinated | 9,908 | 776,645 | 26 | 0.3 | 0.12 (0.06, 0.24) | 88% |
| BNT162b2/Pfizer | 3,883 | 305,677 | 11 | 0.4 | 0.13 (0.05, 0.30) | 87% |
| mRNA-1273/Moderna | 5,556 | 436,890 | 13 | 0.3 | 0.11 (0.05, 0.24) | 89% |
| Ad26.COV2.S/Janssen | 469 | 34,078 | 2 | 0.6 | 0.20 (0.05, 0.90) | 80% |

| June 27 - October 2, 2021 – Delta variant period |  |  |  |  |  |  |
| --- | --- | --- | --- | --- | --- | --- |
| Unvaccinated | 2,599 | 269,780 | 216 | 8.0 | Reference | Reference |
| Partially Vaccinated | 507 | 53,889 | 28 | 5.2 | 0.51 (0.34, 0.77) | 49% |
| Fully Vaccinated | 10,592 | 959,167 | 335 | 3.5 | 0.22 (0.16, 0.30) | 78% |
| BNT162b2/Pfizer | 4,211 | 379,184 | 159 | 4.2 | 0.26 (0.19, 0.36) | 74% |
| mRNA-1273/Moderna | 5,854 | 535,121 | 147 | 2.7 | 0.17 (0.12, 0.24) | 83% |
| Ad26.COV2.S/Janssen | 507 | 44,862 | 29 | 6.5 | 0.38 (0.24, 0.60) | 62% |

CI = confidence interval; \* = Although patients can contribute time to any vaccination status, the N in the first column refers to patients' status at the end of follow-up. Unvaccinated includes patients who never received a vaccine or recipients of a single dose of a vaccine within 14 days of vaccine receipt; partially vaccinated includes patients who were $\geq 14$ days after the first mRNA vaccine dose but $< 14$ days after the 2nd mRNA vaccine dose; fully vaccinated patients includes all patients $\geq 14$ days after the last vaccine dose.
Adjusted for age, days since full vaccination status, female, number of comorbidities, race (white = reference), congregational living (e.g., nursing home, long-term care facility), hemodialysis single pool Kt/V $\geq 1.2$ or peritoneal dialysis weekly Kt/V $\geq 1.7$ , albumin, Hepatitis B seroimmunity defined as hepatitis B surface antibody $\geq 10$ mIU/mL, disability, diabetes, hypertension, peripheral vascular disease, thyroid disease, cancer, county rates for Biden voters and daily rate of COVID infection.

Over the entire study period and compared to unvaccinated patients, the COVID-19 case rate was significantly lower in fully vaccinated than in unvaccinated patients: 2.0 vs 3.8 per 10,000 patient days [adjusted HR 0.15 (0.11, 0.20)](**Table 2**). Vaccine effectiveness was 85% overall, with mRNA-1273/Moderna vaccine having highest vaccine effectiveness at 88%followed by BNT162b2/Pfizer at 83% and Ad26.COV2.S/Janssen at 73%. COVID-19 case rates and vaccine effectiveness during the pre-Delta and Delta variant periods were markedly different. Among all vaccination status groups, COVID-19 case rates increased and vaccine effectiveness against infection decreased during the Delta period. Nonetheless, patients fully vaccinated had lower COVID-19 case rates (3.5 vs. 8.0 per 10,000 patient days [adjusted HR 0.22 (0.16, 0.30)]) and 78% vaccine effectiveness compared to unvaccinated patients during the Delta dominant period, with mRNA-1273/Moderna vaccine had highest vaccine effectiveness of 83%. In the subset analysis of patients vaccinated during the Delta variant dominant period, compared to unvaccinated patients, those fully vaccinated with mRNA-1273/Moderna experienced lowest case rate per 10,000 patient days and highest vaccine effectiveness against COVID-19 infection. (**Table S1**)

### Vaccination status and Vaccine Effectiveness against COVID-19 related hospitalization or death

There were 361 COVID-19 related hospitalizations/deaths during the study period: 188 (52%) among unvaccinated, 41 (11%) among partially vaccinated, and 132 (37%) among fully vaccinated patients (**Table 3**). There were 24 COVID-19 related deaths (mRNA-1273/Moderna N=8; BNT162b2/Pfizer N=14; A26.COV2.S/Janssen N=2), with 19 (79%) occurring while hospitalized and 5 (21%) occurring while patient was at home. Nine (38%) patients having COVID-related death were considered immunocompromised. During the overall study period, the incidence of COVID-19 related hospitalization/death, was 1.35 per 10,000 patient days among unvaccinated and 0.43 per 10,000 patient-days among vaccinated patients [adjusted hazard ratio 0.19 (0.12, 0.30)]. For fully vaccinated patients the vaccine effectiveness was 81% overall (83% with mRNA-1273/Moderna, 75% with BNT162b2/Pfizer and 66% with A26.COV2.S/Janssen vaccine). (**Table 3**).

**Table 3.** COVID-related hospitalization/death rates per 10,000 patient days and vaccine effectiveness between February 1 and October 2, 2021.

|  | Last Vaccination Status* | Person Days at Risk | Events within 30 days COVID-19 diagnosis | Rate per 10,000 days | Adjusted Hazard Ratio (95% CI) | Vaccine Effectiveness (95% CI) |
| --- | --- | --- | --- | --- | --- | --- |
| February 1 – October 2, 2021 |  |  |  |  |  |  |
| Unvaccinated | 3,794 | 1,396,927 | 188 | 1.35 | Reference | Reference |
| Partially Vaccinated | 733 | 353,641 | 41 | 1.16 | 1.09 (0.77, 1.54) | -9%<sup>†</sup> |
| Fully Vaccinated | 11,191 | 1,819,659 | 132 | 0.73 | 0.19 (0.12, 0.30) | 81% |
| BNT162b2/Pfizer | 4,490 | 722,599 | 70 | 0.97 | 0.25 (0.16, 0.40) | 75% |
| mRNA-1273/Moderna | 6,156 | 1,014,171 | 51 | 0.50 | 0.14 (0.08, 0.22) | 83% |
| Ad26.COV2.S/Janssen | 545 | 82,889 | 11 | 1.33 | 0.34 (0.17, 0.68) | 66% |
| February 1- June 26, 2021 – pre-Delta variant period |  |  |  |  |  |  |
| Unvaccinated | 3,809 | 949,295 | 90 | 0.95 | Reference | Reference |
| Partially Vaccinated | 664 | 282,118 | 28 | 0.99 | 1.21 (0.77, 1.89) | -21% |
| Fully Vaccinated | 9,908 | 777,234 | 14 | 0.18 | 0.16 (0.06, 0.42) | 84% |
| BNT162b2/Pfizer | 3,883 | 305,822 | 8 | 0.26 | 0.23 (0.08, 0.68) | 77% |
| mRNA-1273/Moderna | 5,556 | 437,334 | 4 | 0.09 | 0.08 (0.02, 0.29) | 92% |
| Ad26.COV2.S/Janssen | 469 | 34,078 | 2 | 0.59 | 0.54 (0.11, 2.52) | 46% |
| June 27 - October 2, 2021 – Delta variant period |  |  |  |  |  |  |
| Unvaccinated | 2,599 | 274,965 | 98 | 3.56 | Reference | Reference |
| Partially Vaccinated | 507 | 45,608 | 13 | 2.85 | 0.80 (0.44, 1.43) | 20% |
| Fully Vaccinated | 10,592 | 975,676 | 121 | 1.24 | 0.13 (0.08, 0.23) | 87% |
| BNT162b2/Pfizer | 4,211 | 386,000 | 64 | 1.66 | 0.17 (0.10, 0.31) | 83% |
| mRNA-1273/Moderna | 5,854 | 543,216 | 48 | 0.88 | 0.09 (0.05, 0.17) | 91% |
| Ad26.COV2.S/Janssen | 507 | 46,460 | 9 | 1.94 | 0.19 (0.08, 0.43) | 81% |

CI = confidence interval; \* = Although patients can contribute time to any vaccination status, the N in the first column refers to patients' status at the end of follow-up. Unvaccinated includes patients who never received a vaccine or recipients of a single dose of a vaccine within 14 days of vaccine receipt; partially vaccinated includes patients who were $\geq 14$ days after the first mRNA vaccine dose but $< 14$ days after the 2nd mRNA vaccine dose; fully vaccinated patients includes all patients $\geq 14$ days after the last vaccine dose.; † = patient hospitalized for COVID-19 prior to receipt of second mRNA vaccine.
Adjusted for age, days since full vaccination status, female, number of comorbidities, race (white = reference), congregate living (e.g., nursing home, long-term care facility), hemodialysis single pool Kt/V $\geq 1.2$ or peritoneal dialysis weekly Kt/V $\geq 1.7$ , albumin, Hepatitis B seroimmunity defined as hepatitis B surface antibody $\geq 10$ mIU/mL, disability, diabetes, hypertension, peripheral vascular disease, thyroid disease, cancer, county rates for Biden voters and daily rate of COVID infection.

Among all vaccination status groups, both COVID-19 case rates and vaccine effectiveness against COVID-19 related hospitalization/death worsened during the Delta period. In the model comparing unvaccinated, partially vaccinated and fully vaccinated patients, fully vaccinated patients had lowest case rate per 10,000 patient days and adjusted HR for COVID-19 infection and related hospitalization/death in both pre-Delta and Delta dominant periods. In the model comparing unvaccinated, partially vaccinated and each vaccine, patients fully vaccinated with mRNA-1273/Moderna experienced lowest case rate per 10,000 patient days and highest vaccine effectiveness against COVID-19 related hospitalization/death in both pre-Delta and Delta variant dominant periods.

In the pre-Delta period, factors associated lower risk of COVID-19 related hospitalization or death included being vaccinated with BNT162b2/Pfizer (HR [95% CI]: 0.22 [0.07, 0.67]) and mRNA-1273/Moderna; (HR [95% CI]: 0.08 [0.02, 0.30]), whereas being female (HR [95% CI]: 1.55 [1.10, 2.20]) and residence in a congregate setting (HR [95% CI]: 2.21 [1.37, 3.28]) were associated with higher risk (**Table S2)**. During the Delta variant dominant period, all vaccines were associated with lower risk of COVID-19 related hospitalization or death as well as higher serum albumin concentration and hepatitis B surface antibody ≥ 10 mIU/ml, signifying prior seroresponse to hepatitis B vaccine. Similar to the pre-Delta period, congregate living remained a significant risk factor for COVID-19 related hospitalization/death (HR [95% CI]: 1.97 [1.44, 2.70]) in the Delta dominant period. Diabetes as a comorbidity as well as increasing days since full vaccination status were associated with higher risk.

In the subset analysis of patients vaccinated during the Delta variant dominant period, compared to unvaccinated patients, those fully vaccinated experienced lowest case rate per 10,000 patient days and highest vaccine effectiveness against COVID-19 related hospitalization/death. (**Table S1**) Congregate living was the only factor associated with COVID-19 related hospitalization/death in patients vaccinated during the Delta variant dominant period. (**Table S3**)

### Anti-spike IgG level and breakthrough COVID-19

Anti-spike IgG levels were available in 2,451 (22%) fully vaccinated patients over the study period. At each anti-spike IgG threshold evaluated, the lower anti-spike IgG level was associated with higher case rates per 10,000 days and adjusted hazard ratios for both infection (**Table 4**) and COVID-related hospitalization or death (**Table 5**).

**Table 4.** Association of peri-infection anti-spike IgG values with risk for COVID diagnosis.

| Anti-spike IgG threshold | Person Days at Risk | COVID-19 diagnoses | Rate per 10,000 days | Adjusted Hazard Ratio (95% CI) |
| --- | --- | --- | --- | --- |
| <1 | 98,620 | 40 | 4.06 | 3.30 (2.10,5.19) |
| ≥1 | 314,859 | 40 | 1.27 | Reference |
| Total | 413,479 | 80 | 1.93 |  |
| <2 | 123,962 | 52 | 4.19 | 4.66 (2.90,7.49) |
| ≥2 | 289,517 | 28 | 0.97 | Reference |
| Total | 413,479 | 80 | 1.93 |  |
| <7 | 179,341 | 61 | 3.40 | 4.53 (2.68,7.67) |
| ≥7 | 234,138 | 19 | 0.81 | Reference |
| Total | 413,479 | 80 | 1.93 |  |
| <10 | 194,847 | 63 | 3.23 | 4.54 (2.63,7.84) |
| ≥10 | 218,632 | 17 | 0.78 | Reference |
| Total | 413,479 | 80 | 1.93 |  |

Adjusted for age, days since full vaccination status, female, number of comorbidities, race (white = reference), congregate living (e.g., nursing home, long-term care facility), hemodialysis single pool Kt/V ≥ 1.2 or peritoneal dialysis weekly Kt/V ≥ 1.7, albumin, Hepatitis B seroimmunity defined as hepatitis B surface antibody ≥ 10 mIU/mL, disability, diabetes, hypertension, peripheral vascular disease, thyroid disease, cancer, county rates for Biden voters and daily rate of COVID infection.

**Table 5.**
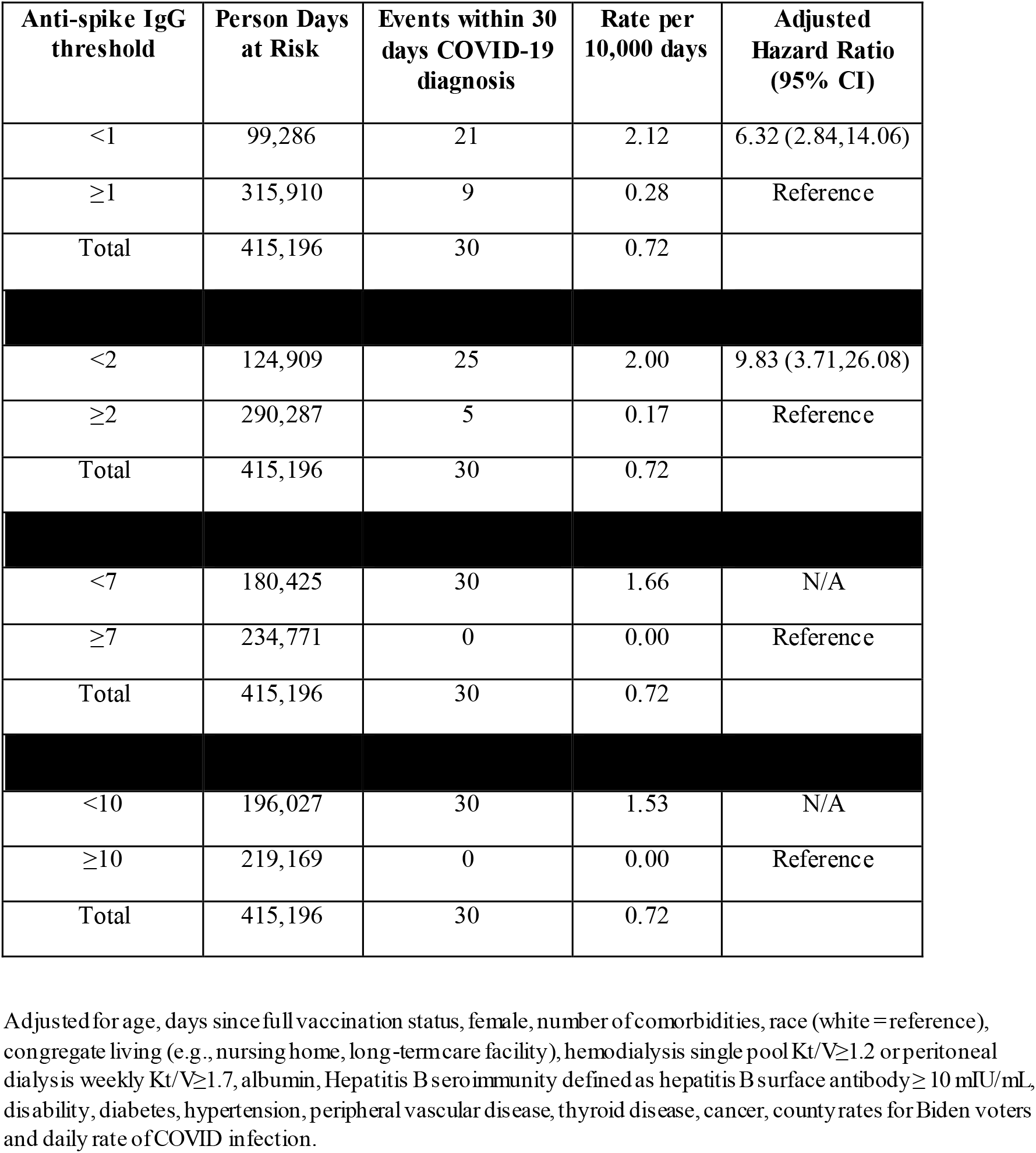
Association of peri-infection anti-spike IgG values with risk for COVID-related hospitalization or death.

There were 80 breakthrough infections (BNT162b2/Pfizer N=32, mRNA-1273/Moderna N=28, and Ad26.COV2.S/Janssen N=20) with the level most proximate to diagnosis measured a median (IQR) of 16 (8, 27) days prior to COVID-19 diagnosis. The frequency of COVID-19 cases requiring hospitalization across anti-spike IgG levels is shown in **Figure 1**. Half of breakthrough cases and the majority of COVID-related hospitalizations (20 of 29) occurred when the anti-spike IgG level was undetectable while an additional 4 hospitalizations and 12 total cases occurred at a level < 2, with the remaining hospitalizations occurring at antibody levels between 1 and 6. There were 8 patients with undetectable anti-spike IgG levels that experienced a COVID-19 related death following a hospitalization event (**Figure 1**).

**Figure 1.**
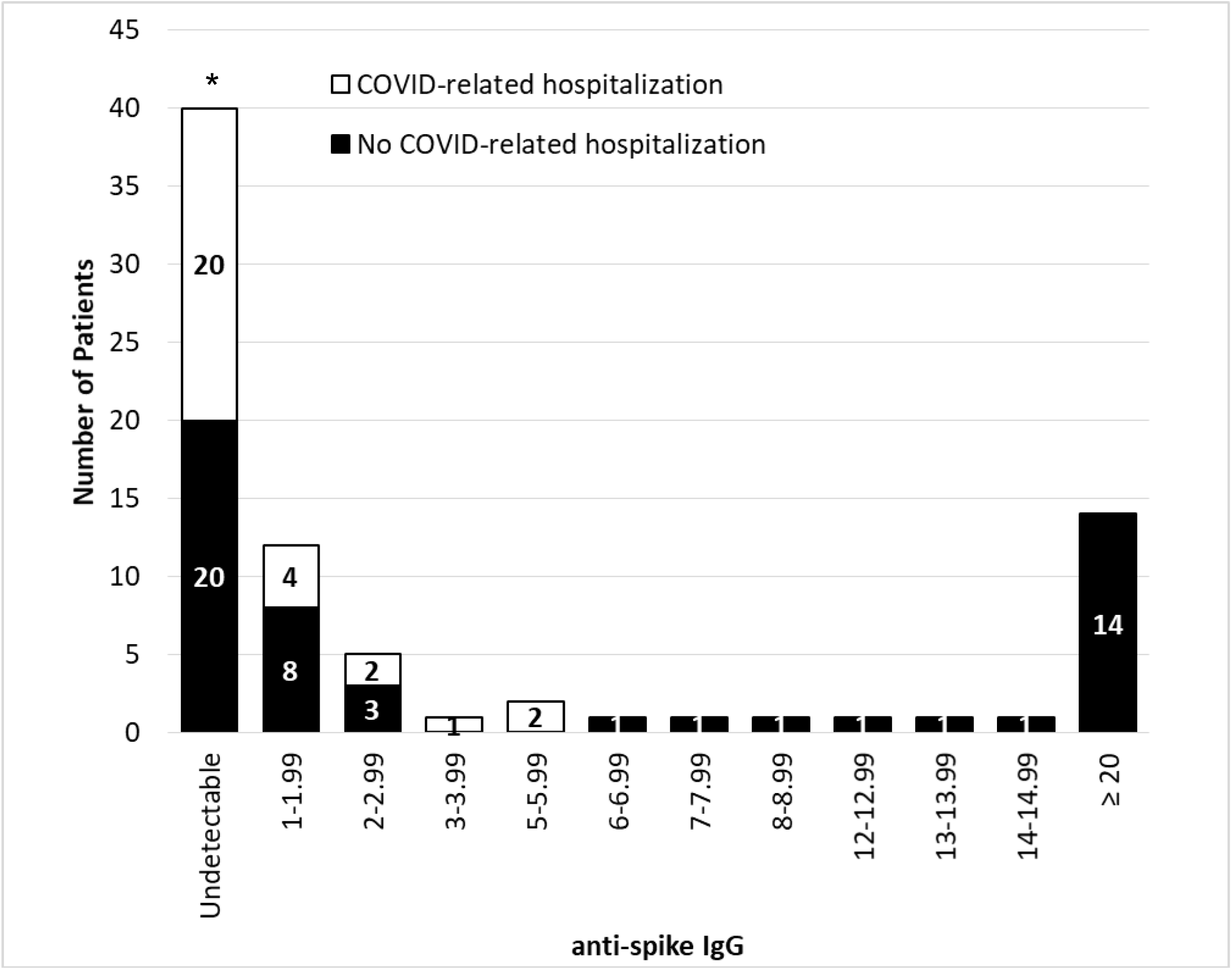
Patient anti-spike IgG value at time of COVID infection diagnosis (N=80) *= COVID-related deaths (n=8); all occurring in patients having COVID-related hospitalization and in those having undetectable anti-spike IgG prior to infection.

## Discussion

SARS-CoV-2 vaccines are highly effective in maintenance dialysis patients, with lower risk for COVID-19 cases and COVID-19 related hospitalization or death among those who are fully vaccinated. Breakthrough COVID-19 cases and COVID-19 related hospitalizations or death among dialysis patients increased when delta variant became dominant, even in those considered fully vaccinated. Although anti-spike IgG antibodies decline over time in vaccinated patients,^8^ vaccine effectiveness in preventing COVID-19 infection and hospitalization/death remains high when compared to unvaccinated patients. Overall, vaccine effectiveness during the Delta dominant period was 10–18% lower than that observed in the pre-Delta period for COVID-19 infection. For COVID-related hospitalization, vaccine effectiveness during presumed Delta variant cases remained high when compared to unvaccinated patients.

Among vaccine types there were marked differences. Overall and during each time period evaluated, vaccine effectiveness for COVID-related hospitalization or death was highest with mRNA-1273/Moderna, then BNT162b2/Pfizer and finally Ad26.COV2.S/Janssen when compared to those unvaccinated. In our study population Ad26.COV2.S/Janssen vaccine was not only associated with lower antibody response,^5, 8^ but likely was associated with higher breakthrough and COVID-19 related hospitalization rates than the mRNA vaccines, particularly when compared to the mRNA-1273/Moderna vaccine.

While the CDC did not specifically designate dialysis patients as immunocompromised persons who should receive routine administration of a third COVID-19 mRNA vaccine dose, they cited dialysis patients as a possible immunocompromised group where clinical opinion is important.^16^ Similar to the general population, the CDC recommends that maintenance dialysis patients receive a booster dose six months or later after full vaccination with mRNA vaccine or a booster of the Ad26.COV2.S/Janssen vaccine two months after initial vaccination. In our results, 50% of breakthrough cases occurred within 4.5 months of full vaccination and 90% of cases occurred within six months post full vaccination status, consistent with a marked increase between 4.5 and 6 months. Consistent with this, increasing time (days) since full vaccination status was associated with increased risk for COVID-19 related hospitalization or death, especially during the Delta period where most patients were several months beyond attaining full vaccination status.

Recognizing that not all maintenance dialysis patients produce anti-spike IgG antibodies to the same degree and that antibodies decline over time, the timing of third dose or a booster should be not arbitrarily be based on time. Since anti-spike IgG antibody titers correlate with COVID neutralizing titers and clinical efficacy,^17, 18^ many clinicians associate detectable antibodies with clinical protection. Presently the CDC does not recommend using COVID-19 antibody testing to guide clinical decision-making.^19^ However, adopting a test and treat approach with routine measurement of anti-spike IgG levels followed by additional doses of vaccine as needed to maintain adequate antibody levels is warranted. Relevantly, we observed high frequency of COVID-19 breakthrough cases and more frequent hospitalization among those with low antibody levels (<7). Recently Anand et al. reported that anti-spike IgG value < 10 were associated with higher odds for breakthrough infection.^15^ In our results (**Tables 3 and 4)**, 79% breakthrough infections occurred in patients having anti-spike IgG value < 10. Additionally, all COVID-related hospitalizations occurred at anti-spike IgG < 6 and COVID-related deaths when antibody levels were undetectable (**Figure 1**). This approach, where vaccine administration is predicated on antibody levels, has been well-demonstrated with hepatitis B vaccination among dialysis patients.^20^

Study strengths include the national population of a mid-size dialysis provider in the US with real world clinical outcomes. However, there are limitations associated with this study. Due to the observational design, residual biases (e.g., misclassification of vaccine exposure in patients vaccinated outside the clinic, inability to identify all asymptomatic infections) and confounding may exist. Randomized clinical trials comparing individual COVID-19 vaccines head-to-head are not likely to be performed in this population. The electronic health records do not contain standardized documentation of COVID-19 symptoms and therefore we could not estimate vaccine effectiveness with regard to mitigating or tempering severity of symptoms. Although the model adjusted for likelihood to follow mask, social distancing and vaccine recommendations throughout the US by adjusting for state and county level presidential election voting records, individual patient actual adherence to those recommendations is not known. Finally, we did not know the specific SARS-CoV-2 variant for each infection and attributed all infections to the Delta variant after June 26, 2021.

In conclusion, SARS-CoV-2 vaccines are effective in maintenance dialysis patients, reducing the risk of both COVID-19 cases and COVID-related hospitalization or death. COVID-19 cases increased during the Delta variant dominant period and current immunosuppression criteria are limited in identifying dialysis patients at highest breakthrough risk. Further research is needed to evaluate the potential utility of antibody titer monitoring to determine patients at highest risk for COVID-19 and to dictate the timing of additional vaccine administration.

## Data Availability

All data produced in the present study are available upon reasonable request to the authors

## Notes

**Support**: This report was supported by Dialysis Clinic, Inc. CMH receives support from ASN KidneyCure’s Ben J. Lipps Research fellowship. CMH’s funder had no role in study design, data collection, reporting, or the decision to submit.

**Financial Disclosure**: Dr. Manley, Mr. Aweh, Dr. Harford, Dr. Johnson and Dr. Lacson Jr are all employees of DCI, where Dr. Johnson is Vice Chair of the Board. Dr. Weiner and Dr. Miskulin receive salary support to their institution from DCI.

### Competing Interest Statement

The authors have declared no competing interest.

### Funding Statement

This report was supported by Dialysis Clinic, Inc. CMH receives support from ASN Kidney Cures Ben J. Lipps Research fellowship. CMH funder had no role in study design, data collection, reporting, or the decision to submit.

### Author Declarations

This study was reviewed and approved by the WCG IRB Work Order 1-1456342-1.

