## supplemental for "SARS-CoV-2 vaccine effectiveness and breakthrough infections in maintenance dialysis patients"

Table S1. COVID-19 infection and COVID-related hospitalization/death rates per 10,000 patient days and vaccine effectiveness between June 27 and October 2, 2021 in patients vaccinated during Delta dominant period.

| | **Last Vaccination Status*** | **Person Days at Risk** | **Events within 30 days COVID-19 diagnosis** | **Rate per 10,000 days** | **Adjusted Hazard Ratio (95% CI)** | **Vaccine Effectiveness (95% CI)** |
|---|---|---|---|---|---|---|
| COVID-19 infection | | | | | | |
| Unvaccinated | 2660 | 274,527 | 217 | 7.9 | Reference | Reference |
| Partially Vaccinated | 467 | 41,541 | 24 | 5.8 | 0.73 (0.47,1.11) | 27% |
| Fully Vaccinated | 636 | 31,411 | 11 | 3.5 | 0.43 (0.24,0.79) | 57% |
| BNT162b2/Pfizer | 321 | 14,325 | 9 | 6.3 | 0.75(0.38,1.47) | 25% |
| mRNA-1273/Moderna | 268 | 14,315 | 1 | 0.7 | 0.09(0.01,0.62) | 91% |
| Ad26.COV2.S/Janssen | 47 | 2,771 | 1 | 3.6 | 0.48(0.07,3.47) | 52% |
| COVID-related hospitalization/death | | | | | | |
| Unvaccinated | 2660 | 282,368 | 86 | 3.05 | Reference | Reference |
| Partially Vaccinated | 467 | 42,216 | 10 | 2.37 | 0.69(0.36,1.34) | 31% |
| Fully Vaccinated | 636 | 31,696 | 4 | 1.26 | 0.37(0.14,1.02) | 63% |
| BNT162b2/Pfizer | 321 | 14,595 | 4 | 2.74 | 0.79(0.29,2.16) | 21% |
| mRNA-1273/Moderna | 268 | 14,325 | 0 | 0.00 | n/a | n/a |
| Ad26.COV2.S/Janssen | 47 | 2,776 | 0 | 0.00 | n/a | n/a |

CI = confidence interval; n/a = not applicable; * = Although patients can contribute time to any vaccination status, the N in the first column refers to patients' status at the end of follow-up. Unvaccinated includes patients who never received a vaccine or recipients of a single dose of a vaccine within 14 days of vaccine receipt; partially vaccinated includes patients who were ≥14 days after the first mRNA vaccine dose but <14 days after the 2nd mRNA vaccine dose; fully vaccinated patients includes all patients ≥14 days after the last vaccine dose.

Adjusted for age, days since full vaccination status, female, number of comorbidities, race (white = reference), congregate living (e.g., nursing home, long-term care facility), hemodialysis single pool Kt/V≥1.2 or peritoneal dialysis weekly Kt/V≥1.7, albumin, Hepatitis B seroimmunity defined as hepatitis B surface antibody ≥ 10 mIU/mL, disability, diabetes, hypertension, peripheral vascular disease, thyroid disease, cancer, county rates for Biden voters and daily rate of COVID infection.

Table S2. Parameter hazard ratios against COVID-19 related hospitalization or death.

| **Parameter** | **pre-Delta variant period** | **Delta variant dominant period** |
|---|---|---|
| | Hazard Ratio (95% Confidence Interval) | Hazard Ratio (95% Confidence Interval) |
| Unvaccinated | Reference | Reference |
| Partially Vaccinated | 1.21 (0.77, 1.90) | 0.79 (0.44, 1.42) |
| Fully vaccinated BNT162b2/Pfizer | **0.22 (0.07, 0.67)** | **0.17 (0.10, 0.31)** |
| Fully vaccinated mRNA-1273/Moderna | **0.08 (0.02, 0.30)** | **0.09 (0.05, 0.17)** |
| Fully vaccinated Ad26.COV2.S/Janssen | 0.55 (0.12, 2.61) | **0.19 (0.08, 0.43)** |
| Weeks since full vaccination status | 1.04 (0.93, 1.17) | **1.17 (1.11, 1.23)** |
| Age (per decade) | 1.29 (0.82, 2.05) | 0.94 (0.86, 1.04) |
| Female | **1.55 (1.10, 2.20)** | 1.08 (0.83, 1.40) |
| Race - Black (reference = White) | 1.03 (0.69, 1.54) | 1.10 (0.82, 1.47) |
| Race - Other (reference = White) | 1.22 (0.75, 1.99) | 1.02 (0.70, 1.48) |
| Congregate living[a] | **2.12 (1.37, 3.28)** | **1.97 (1.44, 2.70)** |
| Albumin g/dL (continuous) | 1.00 (0.69, 1.45) | **0.47 (0.37, 0.60)** |
| Hepatitis B surface antibody ≥ 10 mIU/mL | 0.70 (0.47, 1.03) | **0.67 (0.47, 0.95)** |
| History of failed transplant | 0.49 (0.18, 1.33) | **0.44 (0.19, 0.99)** |
| Number of comorbidities | 0.92 (0.81, 1.04) | 1.08 (1.00, 1.18) |
| Diabetes | 0.97 (0.66, 1.42) | **1.39 (1.03, 1.88)** |
| Hypertension | 1.52 (0.88, 2.62) | 1.16 (0.76, 1.77) |

[a] = Residing in nursing home or long-term care facility; Unvaccinated includes patients who never received a vaccine or recipients of a single dose of a vaccine within 14 days of vaccine receipt; partially vaccinated includes patients who were ≥14 days after the first mRNA vaccine dose but <14 days after the 2nd mRNA vaccine dose; fully vaccinated patients includes all patients ≥14 days after the last vaccine dose.

Table S3. Parameter hazard ratios against COVID-19 related hospitalization or death in patients vaccinated during Delta variant dominant period.

| **Parameter** | Hazard Ratio (95% Confidence Interval) |
|---|---|
| Unvaccinated | Reference |
| Partially Vaccinated | 0.85 (0.44, 1.61) |
| Fully vaccinated BNT162b2/Pfizer | 0.56 (0.10, 3.26) |
| Fully vaccinated mRNA-1273/Moderna | n/a |
| Fully vaccinated Ad26.COV2.S/Janssen | n/a |
| Weeks since full vaccination status | 1.05 (0.79, 1.40) |
| Age (per decade) | 0.91 (0.80, 1.03) |
| Female | 1.40 (0.96, 2.04) |
| Race - Black (reference = White) | 1.16 (0.77, 1.74) |
| Race - Other (reference = White) | 1.22 (0.71, 2.10) |
| Congregate living[a] | **2.01 (1.27, 3.22)** |
| Albumin g/dL (continuous) | 0.75 (0.52, 1.07) |
| Hepatitis B surface antibody ≥ 10 mIU/mL | 0.58 (0.16, 2.07) |
| History of failed transplant | 0.50 (0.16, 1.60) |
| Number of comorbidities | 1.13 (1.00, 1.28) |
| Diabetes | 1.23 (0.81, 1.88) |
| Hypertension | 1.39 (0.74, 2.64) |

[a] = Residing in nursing home or long-term care facility; Unvaccinated includes patients who never received a vaccine or recipients of a single dose of a vaccine within 14 days of vaccine receipt; partially vaccinated includes patients who were ≥14 days after the first mRNA vaccine dose but <14 days after the 2nd mRNA vaccine dose; fully vaccinated patients includes all patients ≥14 days after the last vaccine dose

Figure S1. Cumulative percent COVID-19 breakthrough infections relative to days since full vaccination status overall (panel A) and by vaccine type (panel B)

Panel A

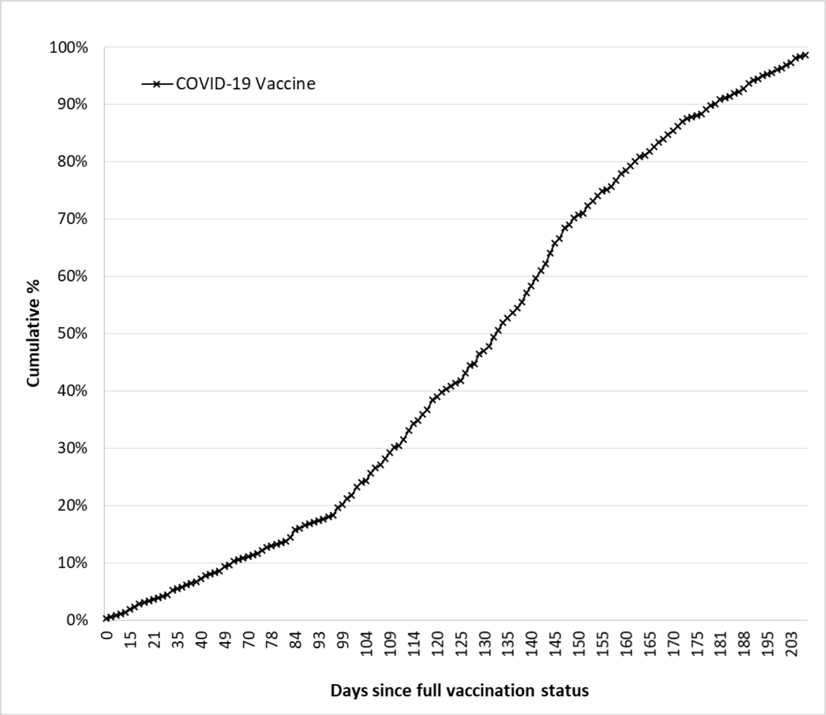


Panel B

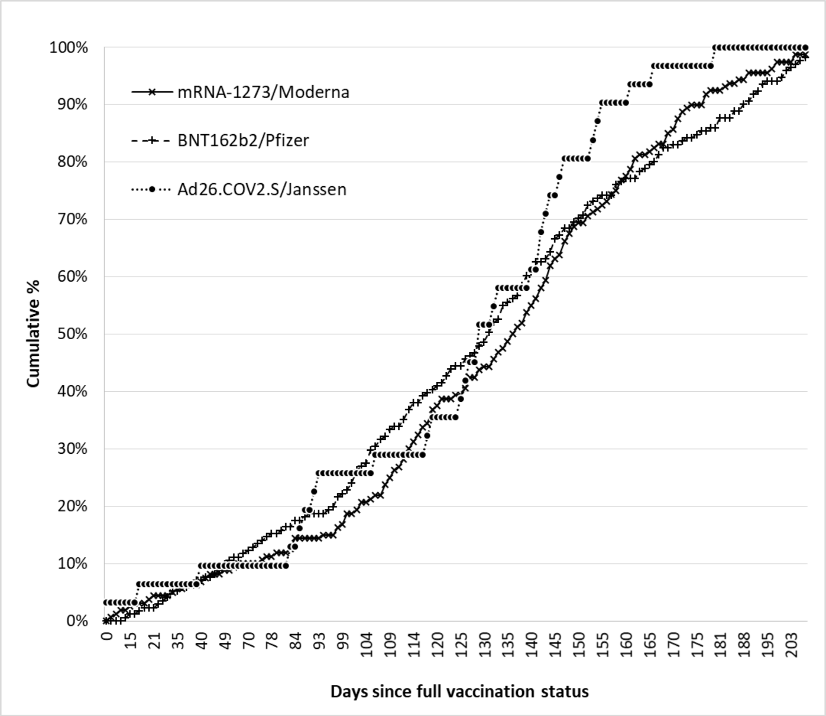
